# Automated, Component-Level Assessment of a National Inpatient Diabetes Guideline: Adherence and Its Association with Hospitalization Complications Across Six Hospitals

**DOI:** 10.64898/2026.09.26.26364069

**Authors:** Shahar Oded, Yuval Shahar, Erez Shalom, Amir Bashkin, Leonard Saiegh, Dror Cantrell, Irit Wirsansky, Muhamad Badarne, Irena Barash, Avivit Cahn, Irit Hochberg

## Abstract

**Objective:** To measure adherence to a national inpatient hyperglycemia guideline at the level of its individual recommendations across a hospital network, to determine whether it changed after the guideline was issued, and to identify which recommendations track subsequent outcomes.

**Materials and Methods:** Retrospective multicenter study of 48,354 admissions among 29,577 adults with diabetes or inpatient hyperglycemia at six Israeli general hospitals, August 2022 to December 2025. Each of 14 recommendations was formalized as a quality assessment temporal pattern over knowledge-based temporal abstractions of the record, scored wherever it applied. Pre–post adherence was compared by Welch *t*-test with false discovery rate correction. Associations with 10 complications were estimated by bias-reduced logistic regression over 24-, 48-, and 72-hour windows, and by inverse probability of treatment weighting contrasting above-with at-or-below-median adherence.

**Results:** Adherence ranged from near-universal admission and medication-management actions to substantial gaps in insulin management (means, 0.15-0.28). Overall adherence rose modestly after issue (0.682 to 0.703, *P <* .001), with marked inter-hospital heterogeneity.After weighting, above-median adherence carried 44% lower odds of 30-day mortality (odds ratio [OR], 0.557; 95% CI, 0.514-0.603), 82% lower odds of hyperglycemia (OR, 0.175) and 52% lower odds of severe hyperglycemia (OR, 0.485), against 20% higher odds of hypoglycemia (OR, 1.204).

**Discussion:** The least-implemented recommendations were largely those whose adherence tracked outcomes: repeated insulin decisions spread across a stay, rather than discrete actions on admission.

**Conclusion:** Guideline adherence can be assessed automatically and per-recommendation at national scale, identifying where fuller implementation would most plausibly matter.

## 1. BACKGROUND AND SIGNIFICANCE

Diabetes mellitus affects approximately 537 million adults worldwide, a figure projected to reach 783 million by 2045^1^. In Israel, the national registry documents a steadily growing patient population and a corresponding burden on the health system^2,3^, concentrated in hospital care: approximately 40% of patients admitted to Israeli general hospitals have diabetes^4^.

In hospital the clinically relevant object is not the diagnosis but the glycemic state during the admission. Hyperglycemia arising in hospital is associated with infection, delayed healing, metabolic decompensation and increased mortality, and occurs in patients with and without documented diabetes. Its counterpart, hypoglycemia, is acute, harmful, and largely iatrogenic—a consequence of the insulin used to correct the first problem. Inpatient glycemic management is therefore a balancing act shaped less by any single decision than by how monitoring, medication management and insulin dosing are carried out repeatedly across a stay. That makes the treatment process itself, not only the patient’s state, worth measuring.

In September 2023 the Israel Endocrine Society, the Israel Diabetes Association and the National Diabetes Council issued a national position paper on inpatient diabetes care^4^, specifying how often glucose should be measured, how oral antidiabetic agents should be handled on admission, when basal and correction insulin should be given, and how to respond to glycemic episodes. Recommendations of this kind are checkable in principle because they are procedural, temporal and conditional: an action is expected, within some window or at some frequency, when a triggering situation holds. Establishing whether they were followed is harder, having conventionally required chart review, which does not scale past modest single-institution samples—too small to separate one ward’s practice from the effect of a guideline, and far too small to observe how practice shifts when a guideline is introduced nationally.

Informatics supplies the machinery to do this automatically. Knowledge-based temporal abstraction and the Temporal Mediator^5^ turn time-stamped clinical data into named interval concepts; Asbru^6,7^ expresses a guideline’s procedural content as a hierarchy of execution plans; and Picard^8^ combines the two to generate real-time recommendations and adherence reports. Deployed in MobiGuide^9^, it raised clinicians’ guideline compliance from 41% to 93%—direct evidence that adherence is malleable, and therefore worth measuring. Retrospective assessment required a third body of knowledge: *quality-assessment* knowledge^10^ re-expresses that procedural content declaratively, as the temporal pattern correct execution would have left in the record, with a membership function grading partial compliance rather than a binary verdict. Such systems have been validated against simulated scenarios^11^ and applied institutionally to long term management at a geriatric hospital^12^.

This study adopts that formulation of adherence but not the systems that introduced it, and departs from them in three respects. In *representation*, procedural and declarative knowledge are encoded in a single schema evaluated by one engine, and the resulting knowledge base is itself a transferable contribution. In *scale*, adherence is measured across six hospitals spanning a national implementation rather than within one institution. In *purpose*, prior work evaluates care *against* the guideline, which serves as the reference standard; here each recommendation is treated as a hypothesis to be tested against what subsequently happened to the patient. Adherence thus becomes an explanatory variable: the question is not whether the guideline was applied correctly, but which of its recommendations demonstrably matter.

## 2. OBJECTIVE

We set out to determine, across a national hospital network spanning the introduction of an inpatient hyperglycemia guideline, how completely each of its individual recommendations was implemented and whether that changed after the guideline was issued; whether adherence is associated with subsequent complications once measured patient characteristics are accounted for; and which individual recommendations carry that association.

## 3. MATERIALS AND METHODS

### 3.1 Study design and data source

We conducted a retrospective multicenter study using electronic health record data from six Israeli general hospitals, drawn from Kineret^13^, the data warehouse of Israel’s largest hospital network, represented in the Observational Medical Outcomes Partnership (OMOP) common data model. The assessed recommendations are those of the national position paper on the treatment of diabetes in hospitalization in a general hospital^4^, issued in September 2023. The pre-intervention period ends immediately before publication (August 2022-July 2023) and the post-intervention period begins a year after it (August 2024-December 2025), the intervening months being excluded to allow for staggered adoption.

Kineret was not mapped to an international coding system at the time of this work, so a dataset-specific concept index—a many-to-one mapping from local codes onto the concepts named in the knowledge base—was curated first. That index is what makes the remainder of the pipeline dataset-agnostic: once it exists, the same knowledge base and engine run unchanged on any source within the same clinical scope. The curated indices are provided as supplementary material, and the curation procedure is described in Supplementary Appendix SI.

### 3.2. Cohort definition

An admission was eligible if it was an inpatient visit of 48 hours to 14 days in a patient aged at least 18 years, with evidence of a genuine overnight stay, no death within 48 hours of admission, no admission to an excluded department, and at least one criterion for diabetes or hyperglycemia. Excluding pediatric and obstetric departments also removes pregnancy-related admissions. Diabetes or hyperglycemia was established by at least one of three non-mutually-exclusive criteria: a documented diabetes diagnosis from a manually curated list of concept identifiers; recurrent hyperglycemia, two glucose measurements ≥180 mg/dL separated by 2 to 24 hours; or extreme hyperglycemia, any glucose measurement ≥250 mg/dL. Hypoglycemia was deliberately not an entry criterion: the cohort is defined by hyperglycemic burden, and hypoglycemia is treated throughout as an outcome rather than as a route into the study population. Full eligibility rules are given in Supplementary Appendix SI.

### 3.3 Abstraction input

The *temporal input* is a long-format table in which each row records one time-stamped clinical fact, with structural tokens inserted to make the admission timeline explicit—notably recurring MEAL tokens, assumed to fall at fixed daily times because meal times are not recorded in the source data. The *context input* holds one row per admission of baseline characteristics from the first 48 hours, from which a modified Charlson comorbidity index (CCI) was computed (Supplementary Appendix SI).

### 3.4. Guideline adherence assessment

Adherence was assessed by knowledge-based temporal abstraction^5^, which converts time-stamped clinical data into named interval concepts and then detects, over those intervals, the temporal pattern that correct execution of each recommendation would have left in the record^10^. Abstraction and assessment are performed in a single pass by our reimplementation of the Temporal Mediator^14^. All domain knowledge is declared externally as XML units validated against a shared schema, so that the clinical semantics and the guideline requirements are stated once, outside the analysis code.

Each recommendation is formalized as a quality-assessment temporal pattern: an *anchor* identifying the clinical situation in which it applies, an *action* that satisfies it, a constraint type,and a membership function grading partial compliance. Applicability is enforced by context—an anchor falling outside the context in which the recommendation holds generates no obligation and is not scored—so that an insulin dose after a meal is required only where glucose is elevated. Recurring recommendations such as routine glucose monitoring are instead evaluated by counting qualifying events within fixed-length windows.

Compliance is graded rather than binary. Each constraint carries a trapezoidal membership function mapping observed timing, frequency, or value to [0,1]: if correction insulin is expected within 4 hours of a hyperglycemic reading and considered missed after 12, insulin given at 8 hours scores 0.5 rather than counting as an outright failure. Three constraint types are used—*time, value*, and *cyclic*—corresponding to the compliance aspects reported below. An unanswered anchor scores 0 where the recommendation requires an action, and 1 where it requires one to be withheld, since there the absence of a response is itself compliant; a recommendation that never became applicable yields no output rather than a zero, so inapplicable recommendations do not depress the score.

Fourteen recommendations were formalized by the first author under the guidance of two coauthors specializing in temporal reasoning and in diabetes care, who reviewed them for clinical fidelity against the source position paper. Scoring is retrospective and offline: the procedure issues no recommendation and returns nothing to the treating team. Thresholds and the full pattern definitions are given in Supplementary Appendices SI and S3.

### 3.5. Outcome definitions

Ten complications were evaluated: 30-day mortality, hyperglycemia, severe hyperglycemia, hypoglycemia, severe hypoglycemia, hyperosmolality, ketoacidosis, infection, inpatient acute kidney injury, and inpatient myocardial infarction. All ten are emitted as abstractions by the same engine, so that outcome ascertainment and adherence assessment draw on a single explicit knowledge base. Thirty-day mortality was taken from linked death records, defined as death between 14 days before and 30 days after the recorded discharge datetime, the backward tolerance capturing in-hospital deaths whose death record precedes the discharge timestamp. Operational definitions are given in Supplementary Appendix S2.

### 3.6. Statistical analysis

The admission is the unit of analysis. Compliance scores were collapsed successively over compliance aspects, calendar days, and finally patterns with equal weight, so that neither frequently-evaluated patterns nor frequently-evaluated days dominate the resulting per-admission score (Supplementary Appendix SI).

Adherence was compared between periods by Welch’s unequal-variance *t-*test, each admission attributed by its admission date. Confidence intervals are unadjusted Welch t-intervals, and *P* values were adjusted across component-level comparisons by the Benjamini-Hochberg false discovery rate (FDR) procedure^15^. Monthly means carry 95% Cis from the within-month standard error, and the temporal trend is an ordinary least-squares fit to those means.

Whole-admission adherence was compared between admissions ending in death within 30 days and those not, and the graded relationship tested by logistic regression on the continuous score, reported as the odds ratio (OR) per 0.1-point increase, then adjusted for age, sex, modified CCI, and hospital with standard errors clustered on patient identity. Both are descriptive, since whole-admission adherence does not strictly precede the outcome.

To identify which individual recommendations were associated with each complication, one model was fitted per complication at cutoffs of 24, 48, and 72 hours. Each row is an admission complication pair, labelled positive if the complication first occurred at or after the cutoff and negative if it never occurred; admissions in which it occurred *before* the cutoff were excluded, since a pre-existing condition cannot be attributed to subsequent in-stay care, and adherence and covariates were computed only up to the cutoff. Models were fitted by bias-reduced (Firth penalized-likelihood) logistic regression^16^, unpenalized maximum likelihood having failed to converge under separation for most complications. Correlated predictors were pruned and the remainder standardized, and a predictor was considered robust only if it remained significant with consistent direction across all three windows. To avoid mechanical circularity, routine glucose monitoring was excluded from the glycemic models, since glucose measurement is itself the mechanism by which those events are detected.

Because these models estimate conditional predictive associations rather than treatment effects, overall adherence and the recurrent components were re-evaluated by inverse probability of treatment weighting, with adherence dichotomized at the median of each outcome’s eligible population. The binary exposure is intrinsic to the design rather than a simplification, since the propensity score^17^ and the standardized mean differences (SMDs) used to verify balance^18^ are two-group constructs undefined for a continuous exposure; retaining continuous adherence in the regressions above therefore makes the two designs complementary. Propensity scores were estimated by bias-reduced logistic regression, converted to stabilized weights and trimmed at the 1st and 99th percentiles. Balance was assessed by the maximum post-weighting SMD, with <0.10 required for an estimate to be interpretable; pairs failing this are reported as non-estimable. Confidence intervals are unadjusted throughout, so an interval excluding the null may accompany a nonsignificant adjusted *P* value. Analyses used Python 3.13 (Supplementary Appendix SI).

## 4. RESULTS

### 4.1. Study cohort

The cohort comprised 48,354 admissions among 29,577 unique adult patients across six Israeli general hospitals, with center-specific sample sizes ranging from 4,478 to 11,024 admissions (Table 1). Only 21.5% of admissions meeting a hyperglycemia criterion carried a documented diabetes diagnosis, indicating that a substantial proportion of the cohort represented clinically significant inpatient hyperglycemia without a recorded diagnosis.

**Table 1:** Characteristics of the multicenter study cohort.

| Characteristic | Value |
| --- | --- |
| Hospitals | 6 |
| Admissions | 48,354 |
| Unique patients | 29,577 |
| Age, mean (median), y | 71.9 (73.3) |
| Male sex | 54.3% |
| Length of stay, median, d | 5.55 |
| 30-day mortality | 9.58% |
| Documented diabetes diagnosis | 35.3% |
| Recurrent hyperglycemia | 74.7% |
| Extreme glucose ($\geq 250$ mg/dL) | 60.2% |
| Hyperglycemia among admissions with diabetes | 50.3% |
| Diabetes diagnosis among admissions with hyperglycemia | 21.5% |
| Pre-intervention cohort | 21,605 admissions<br>17,002 patients |
| Post-intervention cohort | 16,182 admissions<br>11,201 patients |

### 4.2. Adherence and its change following national implementation

Adherence varied sharply across recommendations, and the split followed the kind of action required rather than its clinical weight (Table 2). Single discrete actions on admission were near-universal: SGLT2 discontinuation was complete (1.00) and creatinine measurement almost so (0.98). Recommendations calling for repeated insulin decisions across the stay were not: insulin following hyperglycemia scored 0.15, the two routine basal dosing components 0.21 and 0.23, and bolus reduction after hypoglycemia 0.28—all with a median of zero, meaning the expected action was absent altogether in most eligible admissions. Monitoring and admission medication management fell between these extremes (0.54-0.88). Adherence was largely independent of whether diabetes had been formally documented (0.669 vs 0.661 among admissions with hyperglycemia in the first 48 hours), a difference reaching significance only by virtue of the sample size.

**Table 2:** Adherence to each assessed inpatient diabetes guideline recommendation over the whole study period, and its change following national implementation. Scores range from 0 (nonadherence) to 1 (complete adherence). Δ is the absolute difference in mean adherence between the post- and pre-intervention periods, with unadjusted 95% Cis; *P* values were adjusted across component-level comparisons using the Benjamini-Hochberg false discovery rate procedure.

| Recommendation | Mean | Median | Before | After | $\Delta$ (95% CI) | $P$ |
| --- | --- | --- | --- | --- | --- | --- |
| Admission glucose monitoring | 0.85 | 1.00 | 0.78 | 0.93 | +0.15 (0.145 to 0.154) | < .001 |
| Routine glucose monitoring | 0.88 | 0.98 | 0.83 | 0.94 | +0.11 (0.105 to 0.113) | < .001 |
| Glucose monitoring during corticosteroid therapy | 0.54 | 0.50 | 0.51 | 0.58 | +0.07 (0.062 to 0.087) | < .001 |
| BMI measurement on admission | 0.87 | 1.00 | 0.86 | 0.89 | +0.03 (0.015 to 0.029) | < .001 |
| Creatinine measurement on admission | 0.98 | 1.00 | 0.98 | 0.99 | +0.01 (0.008 to 0.014) | < .001 |
| Antidiabetic discontinuation on admission | 0.66 | 0.86 | 0.62 | 0.75 | +0.13 (0.075 to 0.192) | < .001 |
| SGLT2 discontinuation on admission | 1.00 | 1.00 | 1.00 | 1.00 | 0.00 (–) | – |
| Basal insulin continuation on admission | 0.66 | 1.00 | 0.63 | 0.67 | +0.04 (0.002 to 0.089) | .054 |
| Insulin administration after hyperglycemia | 0.15 | 0.00 | 0.14 | 0.15 | +0.01 (0.009 to 0.020) | < .001 |
| Antidiabetic discontinuation after hypoglycemia | 0.89 | 1.00 | 0.91 | 0.89 | –0.02 (–0.197 to 0.158) | .818 |
| Basal dose reduction after hypoglycemia | 0.87 | 1.00 | 0.87 | 0.88 | +0.01 (–0.015 to 0.034) | .497 |
| Bolus dose reduction after hypoglycemia | 0.28 | 0.00 | 0.33 | 0.26 | –0.07 (–0.158 to 0.029) | .210 |
| Routine basal dosage (frequency) | 0.23 | 0.00 | 0.21 | 0.24 | +0.03 (0.027 to 0.043) | < .001 |
| Routine basal dosage (dose) | 0.21 | 0.00 | 0.19 | 0.22 | +0.03 (0.021 to 0.036) | < .001 |

Overall adherence rose from 0.682 before the guideline to 0.703 after (*P <* .001; Figure 1), with monthly scores trending upward across the period. The absolute change is small, and its significance reflects the number of admissions rather than the size of the shift. Component-level changes were uneven (Table 2). Glucose monitoring improved most—on admission from 0.78 to 0.93—and discontinuation of high-hypoglycemia-risk antidiabetic agents on admission rose from 0.62 to 0.75. Gains in the insulin-management components were significant but marginal (+0.01 to +0.03), leaving them close to their already low baselines, while dose reduction after hypoglycemia did not change and SGLT2 discontinuation was already complete in both periods. Center-level trajectories ranged from improvement through no measurable change to modest decreases, so the pooled increase conceals substantial inter-hospital heterogeneity.

**Figure 1:**
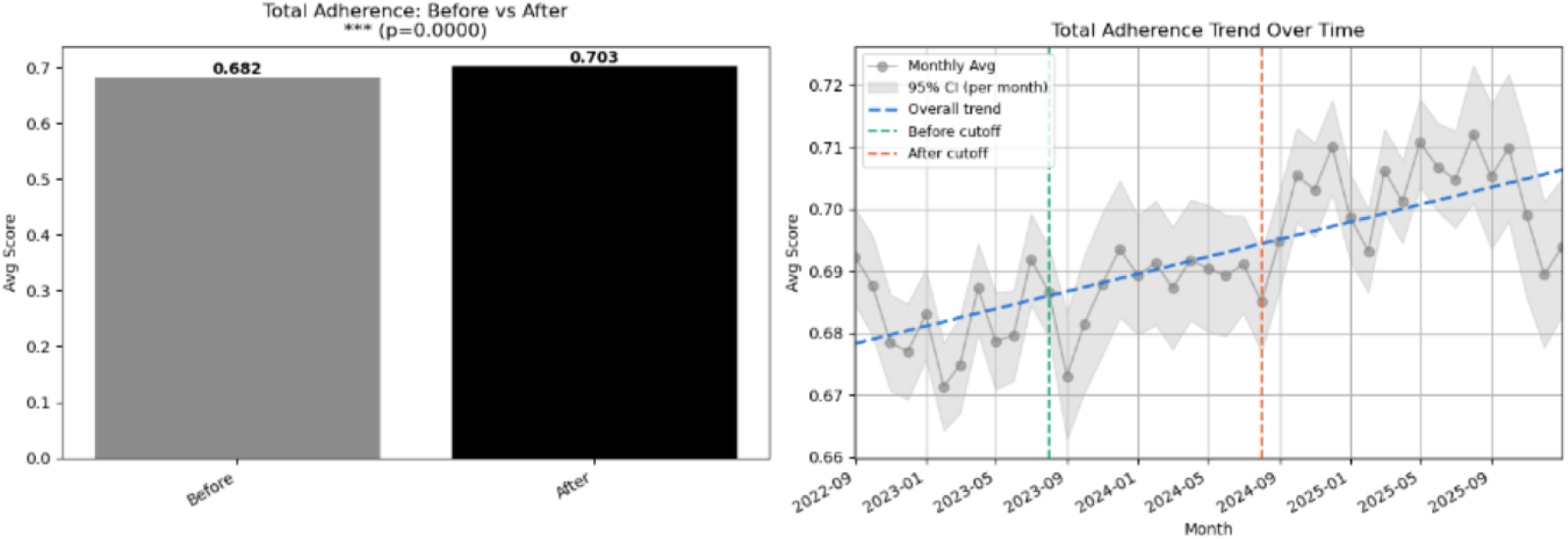
Changes in overall guideline adherence across the study period. **Left:** mean overall adherence during the predefined pre-intervention (August 2022– July 2023) and post-intervention (August 2024 – December 2025) periods. **Right:** monthly mean overall adherence with 95% Cis. Vertical dotted lines delimit the transition period excluded from the primary pre– post comparison; the dashed line indicates the overall temporal trend. *Alt text:* Bar chart and monthly timeline plot illustrating overall guideline adherence before and after September 2023.

### 4.3. Adherence and clinical outcomes

Higher adherence was associated with lower 30-day mortality (Figure 2). Survivors had a higher mean adherence score than those who died (0.693 vs 0.652, *P <* .001), and mortality fell across adherence strata from 11% to roughly 3%. On the continuous score, each 0.1-point increase carried 16.8% lower odds of death (OR, 0.832; 95% CI, 0.811-0.853), so the association is graded rather than threshold-like, and adjusting for age, sex, comorbidity burden and hospital left it intact (adjusted OR, 0.757; 95% CT, 0.727-0.788, per 1-SD increase). Adherence to the insulin-management recommendations showed negligible correlation with subsequent hypoglycemic burden, for both hypoglycemia (*r* = 0.052) and severe hypoglycemia (*r* = 0.056; *n* = 40,719).

**Figure 2:**
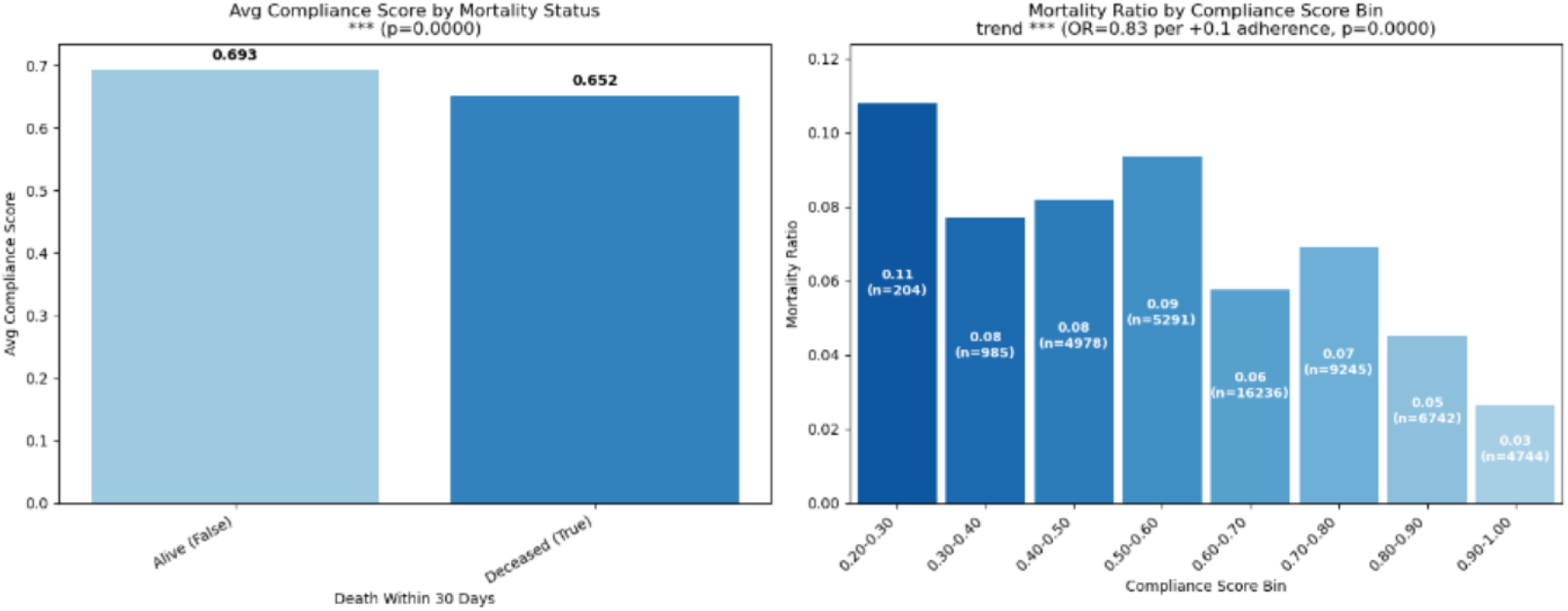
Association between overall guideline adherence and 30-day mortality. **Left:** mean overall adherence among patients who survived and those who died within 30 days. **Right:** observed 30-day mortality across strata of overall adherence, with the number of admissions shown within each bar; strata containing fewer than 30 admissions are not shown. The trend statistic is from logistic regression on the continuous adherence score, not on the binned values, and is expressed as the odds ratio per 0.1-point increase. *Alt text:* Bar charts displaying average guideline adherence by 30-day mortality status and mortality rates across adherence deciles.

### 4.4. Outcome-associated recommendations

Between 9 and 34 predictors were retained per outcome, but the number surviving all three window definitions ranged from none to 32, tracking event frequency: mortality yielded the most robust predictors and the rarer complications few or none (Supplementary Table S2). For mortality, the model recovered well-established clinical predictors—age, red cell distribution width, body mass index (BMI), dementia, cardiovascular comorbidity—which gives the approach face validity, and several adherence components ranked among the strongest retained predictors (Figure 3). Across all outcome models (Figure 4), insulin following hyperglycemia and admission glucose monitoring recurred most broadly, while basal-dose components appeared in fewer, more outcome-specific models. Direction was not uniform—the same component could carry opposite signs for different endpoints.

**Figure 3:**
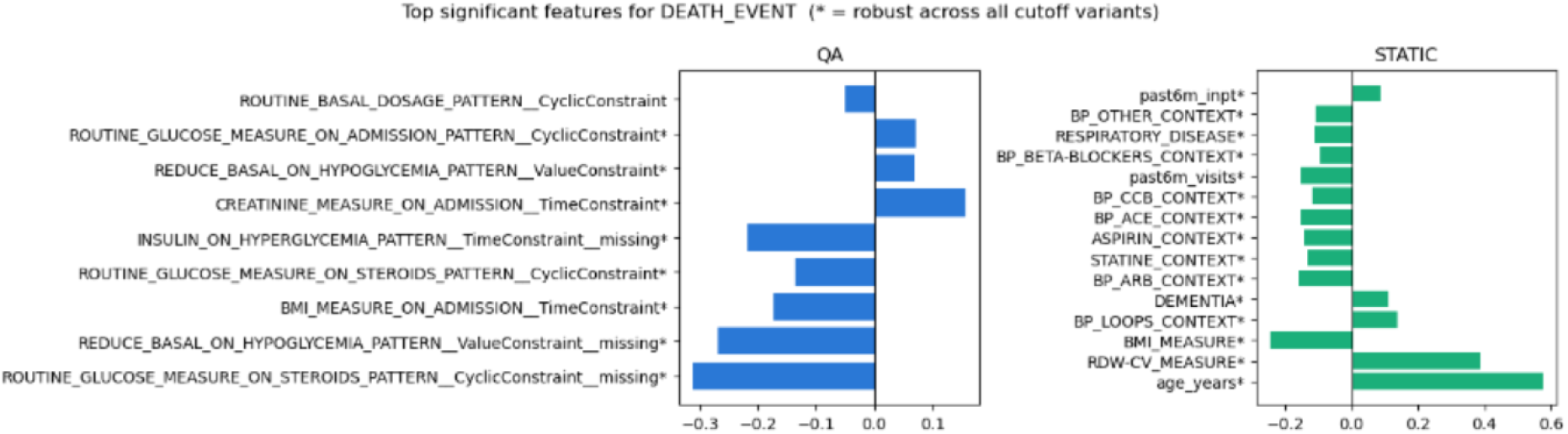
Retained feature profile for 30-day mortality. The left panel shows retained guideline adherence (quality-assessment) features and the right panel retained baseline clinical variables. Bar direction indicates the sign of the regression coefficient, and bar magnitude the relative contribution after false discovery rate correction and correlation pruning. Features marked with an asterisk persisted across all three observation-window definitions. *Alt text:* Horizontal bar plots comparing regression coefficients for quality assessment patterns and baseline clinical variables predicting 30-day mortality.

**Figure 4:**
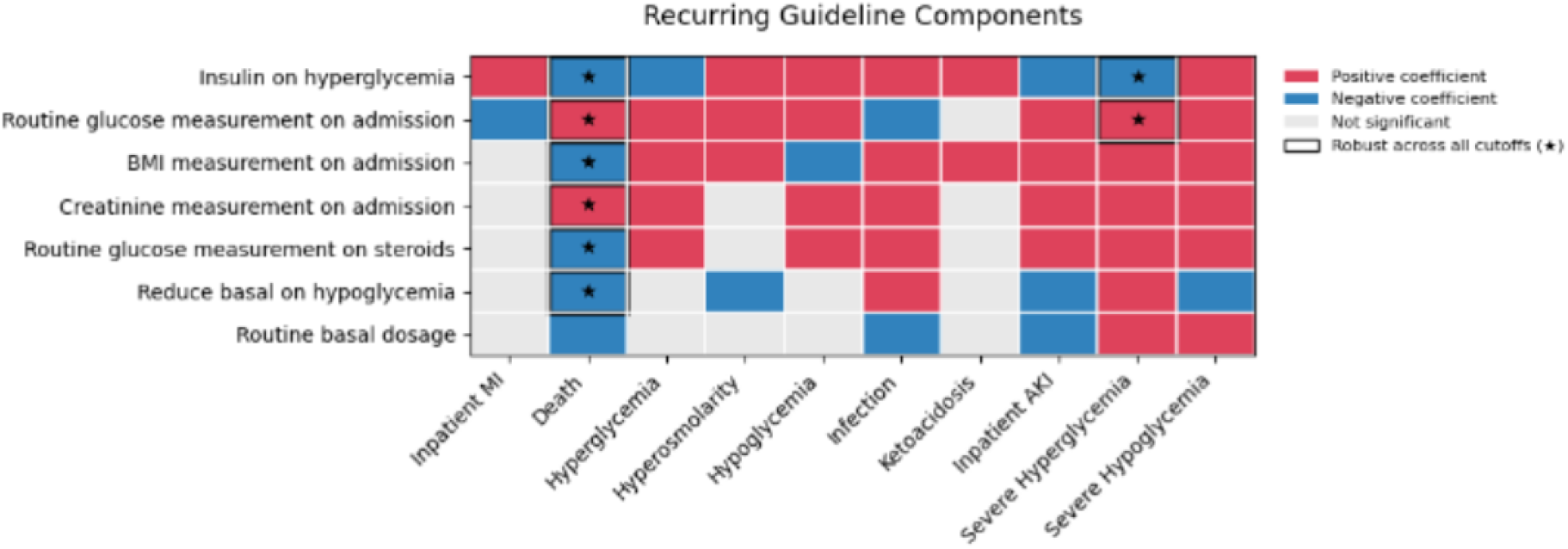
Recurring guideline-adherence components identified by the bias-reduced regression models across clinical outcomes. Rows correspond to guideline recommendations and columns to clinical outcomes. Colored cells indicate statistically significant associations after false discovery rate correction (red, positive coefficient; blue, negative coefficient). Stars denote associations that remained significant with a consistent direction across all three observation-window definitions (24, 48, and 72 hours). Blank cells indicate no statistically significant association. *Alt text:* Heatmap matrix showing recurring guideline adherence components across ten clinical complications with direction of association.

### 4.5. Propensity-weighted evaluation

The overall-adherence analyses achieved satisfactory covariate balance (maximum post-weighting SMD <0.08). Relative to the low-adherence group, above-median adherence carried 44% lower odds of 30-day mortality (OR, 0.557; 95% CI, 0.514-0.603), 82% lower odds of hyperglycemia (OR, 0.175) and 52% lower odds of severe hyperglycemia (OR, 0.485), all adjusted *P <* .001, against 20% higher odds of hypoglycemia (OR, 1.204; 95% CI, 1.090-1.331; Table 3). Nothing survived correction for the remaining five outcomes. The asymmetry between those two directions is the substantive result: the reduction in hyperglycemic burden is roughly ninefold larger on the log-odds scale than the accompanying increase in hypoglycemia, and the increase in *severe* hypoglycemia did not reach significance (OR, 1.129; adjusted *P* = .096).

**Table 3:** Inverse probability of treatment weighting analysis of overall guideline adherence. Adherence was dichotomized at the median of each outcome’s eligible population, so *β* is the log-odds ratio comparing the above-median with the at-or-below-median group and OR = exp(*β*). Confidence intervals are unadjusted; *P* values were adjusted across outcomes using the Benjamini-Hochberg false discovery rate procedure. Balance was assessed by the maximum standardized mean difference (SMD) after weighting. AKI, acute kidney injury; MI, myocardial infarction.

| Outcome | $\beta$ | OR | 95% CI (OR) | Max SMD | $P$ |
| --- | --- | --- | --- | --- | --- |
| Hyperglycemia | −1.742 | 0.175 | 0.154 to 0.199 | 0.019 | < .001 |
| Severe hyperglycemia | −0.724 | 0.485 | 0.450 to 0.523 | 0.012 | < .001 |
| 30-day mortality | −0.586 | 0.557 | 0.514 to 0.603 | 0.020 | < .001 |
| Hypoglycemia | 0.186 | 1.204 | 1.090 to 1.331 | 0.034 | < .001 |
| Severe hypoglycemia | 0.121 | 1.129 | 1.001 to 1.274 | 0.030 | .096 |
| Hyperosmolarity | 0.106 | 1.112 | 0.652 to 1.896 | 0.020 | .800 |
| Ketoacidosis | 0.116 | 1.123 | 0.561 to 2.246 | 0.021 | .800 |
| Inpatient MI | 0.055 | 1.057 | 0.732 to 1.523 | 0.020 | .800 |
| Infection | −0.025 | 0.975 | 0.809 to 1.175 | 0.020 | .800 |
| Inpatient AKI | 0.018 | 1.018 | 0.888 to 1.168 | 0.022 | .800 |

Only a subset of the recurrent component-level predictors remained significant after weighting, indicating that several predictive associations were largely explained by differences in baseline patient characteristics. A further six significant associations were set aside because weighting failed to achieve acceptable balance (maximum SMD, 0.90–2.46). All six concerned BMI or creatinine measurement on admission, for which adherence is near-universal, so that dichotomizing at the median produces groups too dissimilar to balance; those coefficients were numerically degenerate and are reported as non-estimable.

Eight associations were both significant and adequately balanced (Table 4). Above-median adherence to routine basal insulin dosing, routine glucose monitoring and insulin administration following hyperglycemia each remained associated with lower 30-day mortality. For glycemic outcomes, insulin following hyperglycemia (78% lower odds; OR, 0.220) and routine basal insulin dosing (60% lower odds; OR, 0.402) were associated with substantially lower odds of subsequent severe hyperglycemia, while insulin following hyperglycemia also carried 28% higher odds of hypoglycemia (OR, 1.283)—the same trade-off seen at the level of overall adherence. Adherence to glucose monitoring on admission was associated with a higher recorded incidence of hyperglycemia and severe hyperglycemia; since these outcomes are ascertained from glucose measurements, more frequent early measurement mechanically increases the chance of recording an event, so both associations likely reflect ascertainment rather than harm.

**Table 4:** Inverse probability of treatment weighting analysis of individual guideline recommendations. Rows are the component-outcome pairs that were both statistically significant after false discovery rate correction and adequately balanced after weighting (maximum standardized mean difference <0.10). A further six significant associations, all concerning BMI or creatinine measurement on admission, failed the balance criterion (maximum standardized mean difference, 0.90-2.46) and are not reported. ^†^ Likely to reflect ascertainment rather than harm: the outcome is detected from glucose measurements, so more frequent early measurement increases the opportunity to record an event.

| Outcome | Recommendation | $\beta$ | OR | 95% CI (OR) | *P* |
| --- | --- | --- | --- | --- | --- |
| 30-day mortality | Routine basal dosage | −0.291 | 0.748 | 0.691 to 0.809 | < .001 |
| 30-day mortality | Routine glucose monitoring | −0.139 | 0.870 | 0.809 to 0.937 | < .001 |
| 30-day mortality | Insulin after hyperglycemia | −0.098 | 0.907 | 0.842 to 0.977 | .011 |
| Severe hyperglycemia | Insulin after hyperglycemia | −1.514 | 0.220 | 0.202 to 0.239 | < .001 |
| Severe hyperglycemia | Routine basal dosage | −0.912 | 0.402 | 0.372 to 0.434 | < .001 |
| Severe hyperglycemia | Admission glucose monitoring<sup>†</sup> | 0.752 | 2.121 | 1.974 to 2.280 | < .001 |
| Hyperglycemia | Admission glucose monitoring<sup>†</sup> | 0.980 | 2.664 | 2.389 to 2.971 | < .001 |
| Hypoglycemia | Insulin after hyperglycemia | 0.249 | 1.283 | 1.163 to 1.416 | < .001 |

These are the same components that showed the largest implementation gaps: insulin following hyperglycemia (mean adherence, 0.15) and routine basal insulin dosing (0.21–0.23) are simultaneously the least-implemented recommendations assessed and, after weighting, among those most strongly associated with mortality and with severe hyperglycemia.

## 5. DISCUSSION

### 5.1. Principal findings

Adherence to the national inpatient hyperglycemia recommendations improved after their introduction, and higher adherence was consistently associated with lower mortality and reduced hyperglycemic complications. Adherence was not uniform: near-universal implementation was observed for several admission and medication-management actions, whereas the largest gaps were concentrated in insulin management. This pattern separates recommendations satisfied by a single discrete action, such as ordering a laboratory test or discontinuing a medication on admission, from those requiring repeated insulin decisions across the hospitalization. The improvement was significant but modest, its *P* value reflecting the number of admissions rather than the size of the effect, and it was accompanied by substantial inter-hospital heterogeneity that the data do not explain.

The clearest outcome signal was for 30-day mortality. The propensity-weighted estimate is primary, being the only one whose exposure window—a fixed early-admission period—is guaranteed to precede the outcome. Two descriptive estimates agree in direction: whole-admission adherence adjusted for baseline characteristics, and a graded fall in mortality across the adherence range. A graded association is harder to explain by a simple difference in case mix, though it shares the whole-admission timing concern noted below. Because the three use different exposure definitions, they corroborate direction rather than magnitude.

The glycemic findings show a favorable risk profile. Greater adherence bought a large reduction in hyperglycemic burden—roughly ninefold larger on the log-odds scale than the accompanying rise in hypoglycemia—at the cost of a small increase in mild hypoglycemia and no detectable increase in severe hypoglycemia, which is the balance the recommendations are intended to strike. The same asymmetry appears at component level, where insulin following hyperglycemia tracked lower severe hyperglycemia alongside more hypoglycemia. Since nonsignificance is not evidence of absence, the severe-hypoglycemia result shows no signal of harm at this sample size rather than establishing none exists.

Adherence features ranked among the strongest retained predictors alongside established clinical variables—the practical requirement for treating them as monitorable quality targets. These findings have no direct precedent: the position paper was issued in 2023, and no prior study has measured adherence to it at this scale.

### 5.2. Implications

The most actionable pattern in these results is a convergence. The recommendations showing the largest implementation gaps—insulin following hyperglycemia, and the frequency and dose components of routine basal insulin—are also among those most consistently associated with subsequent outcomes after balancing measured baseline characteristics. The components where practice departs most from the guideline are therefore not peripheral ones, and this identifies them as natural targets for decision support: they are repeated insulin decisions distributed across a hospitalization, precisely the kind of task where a prompt at the point of care can plausibly help, and unlike the admission recommendations they are not already at ceiling.

This should not be read as an attribution of fault. A low adherence score records that the action expected by the guideline is absent from the record, not that the decision was wrong. Ward-level judgment—a patient not eating, an imminent procedure, a deliberate decision to tolerate higher glucose in a frail patient—is frequently the correct reason for a departure, and none of it is recoverable from records collected for clinical rather than research purposes. The measure is a population-level statistic, as is the inter-hospital variation reported here. What the work offers is a measurement instrument—an automated, component-level and transferable way to ask which of a guideline’s recommendations were followed and which track outcomes—rather than a decision-support system.

Nothing restricts the approach to recommendations a guideline already contains. Because each is declared externally as a pattern rather than implemented in analysis code, a practice merely suspected to matter costs no more to evaluate than the patterns scored here. That inverts the question: one could ask which candidate practices earn a place in a guideline, and—more uncomfortably—which current recommendations do not track outcomes at all. Such candidates would be hypotheses for prospective evaluation, and a wide search would need more conservative multiplicity correction than applied here.

### 5.3. Limitations

The design is observational, and none of the estimates should be read as causal effects. Residual confounding remains throughout: weighting balances only measured covariates, and unmeasured factors—real-time illness severity above all, and decisions to limit or de-escalate treatment— cannot be excluded. Direction was not uniform across outcomes, reflecting differences in indication and applicability.

The mortality analyses divide by exposure definition. The component-level and propensity-weighted models score adherence over a fixed early-admission window, so exposure strictly precedes outcome. The adjusted estimate instead averages adherence over the whole admission and is descriptive: because 30-day mortality is predominantly an end-of-admission event, and guideline-concordant management is frequently de-escalated in patients on a deteriorating trajectory, that average incorporates the period immediately preceding death, where adherence may reflect the course of care rather than determine it. Dichotomizing adherence at the median, as the weighting requires, discards dose-response information and costs power.

Adherence and outcomes were ascertained during the same hospitalization, and detecting glycemic events depends on glucose being measured; routine glucose monitoring was therefore excluded from the glycemic models, though residual detection effects may remain for other monitoring recommendations. The pre-post comparison is uncontrolled, so secular trends in inpatient care cannot be separated from the implementation itself, and the non-mortality models do not account for the competing risk of death, which biases those endpoints conservatively. The findings come from adult inpatients in Israeli general hospitals under a single national guideline, so the adherence levels observed and the components that proved most consequential may differ elsewhere. Finally, the data are assembled from hospital admissions rather than from a longitudinal record, so chronic diagnoses managed in the community may be incompletely captured; ascertaining eligibility and glycemic outcomes from glucose measurements rather than diagnosis codes limits the consequences, but the comorbidity score and the proportion of hyperglycemic admissions carrying a documented diagnosis are both likely understated.

## 6. CONCLUSION

Adherence to a national inpatient hyperglycemia guideline can be measured automatically, at the level of its individual recommendations, across an entire hospital network. Applying that measurement to 48,354 admissions in six Israeli general hospitals shows that adherence improved after the guideline was issued, that the improvement was modest and unevenly distributed across both recommendations and centers, and that adherence is associated with lower 30-day mortality and a lower burden of hyperglycemic complications once measured baseline differences are balanced. The recommendations least well implemented are largely the same ones whose adherence tracks subsequent outcomes: the repeated insulin decisions distributed across a hospitalization, rather than the single discrete actions taken on admission. That convergence, together with the favorable shift in glycemic risk, is the evidence a guideline needs to justify implementing it more completely. These are associations rather than treatment effects, and they describe how a guideline was adopted rather than how well any clinician practised. What the work establishes is that the question is now answerable of an entire health system’s records rather than of a sample small enough to review by hand.

## Data Availability

All code, the knowledge base of temporal-abstraction and quality-assessment definitions, and analysis notebooks are publicly available at https://github.com/shaharoded/Mediator and https://github.com/shaharoded/Kineret-ETL[cite: 1]. The curated concept indices are provided as supplementary material[cite: 1]. The underlying patient-level electronic health record data cannot be shared as they comprise identifiable clinical records governed by institutional data agreements and Helsinki committee approvals under which they were obtained; all analyses were conducted inside a secure hospital computing environment and exported as aggregated statistics[cite: 1].

https://github.com/shaharoded/Kineret-ETL

## AUTHOR CONTRIBUTIONS

**Shahar Oded:** conceptualization, methodology, software, validation, formal analysis, data curation, visualization, writing—original draft. **Yuval Shahar:** conceptualization, methodology, supervision, funding acquisition, writing—review and editing. **Erez Shalom:** resources,project administration, writing—review and editing. **Amir Bashkin, Leonard Saiegh, Dror Cantrell, Irit Wirsansky, Muhamad Badarne**, and **Irena Barash:** investigation, resources, writing—review and editing. **Avivit Cahn:** conceptualization, investigation, writing—review and editing. **Irit Hochberg:** conceptualization, methodology, validation, resources, supervision, writing—review and editing.

## SUPPLEMENTARY MATERIAL

Supplementary material is available at *Journal of the American Medical Informatics Association* online. Appendix SI details data preparation, the concept index, the modified Charlson comorbidity index, threshold acquisition, score aggregation, and software. Appendix S2 gives the operational definitions of all ten complications. Appendix S3 gives the operational definitions of the fourteen assessed guideline recommendations. Table S2 summarizes the bias-reduced regression models. The curated concept indices are supplied as a separate data file.

## ACKNOWLEDGMENTS

This study would not have been possible without the Kineret project team, who built and maintain the harmonized multicenter data warehouse on which it rests. Transforming the records of a national hospital network into a common data model, and sustaining the infrastructure and governance that make it usable for research, represents a substantial body of work in its own right; the authors are grateful for that effort and for the team’s support in accessing the data and extracting the study cohort. The authors also thank the Helsinki committee coordinators at the participating centers for their assistance throughout the approval process.

Generative AI tools were used as a writing assistant and to support methodological research and code adaptation. Every design choice, analysis step, and interpretation was directed and verified by the corresponding author and, in turn, by the co-authors.

## DATA AVAILABILITY

The abstraction engine, the knowledge base of temporal-abstraction and quality-assessment definitions, and the analysis notebooks are publicly available in the Mediator repository^14^, and the extraction and cohort-construction pipeline in a companion repository^19^. The curated concept indices mapping the source vocabulary onto the concepts named in the knowledge base are provided as supplementary material.

All processing and analysis were performed inside a closed hospital computing environment. The code released here is a replica of that pipeline; no source data, intermediate datasets, or unaggregated outputs leave that environment, and the results reported in this paper are exported aggregate statistics. The underlying patient-level electronic health record data cannot be shared, as it comprises identifiable clinical records governed by the approvals under which it was obtained.

## ETHICS APPROVAL

The study was approved by the institutional Helsinki committee of each participating center under a common protocol (NHR-0152-23): Galilee Medical Center (NHR-0152-23), Hillel Yaffe Medical Center (HYMC-0074-24), Shamir Medical Center (ASF-0117-24), Tzafon Medical Center (POR-0047-24), Barzilai University Medical Center (BRZ-0059-24), and Bnai Zion Medical Center (BNZ-0067-24). Recruitment was approved for up to 30,000 participants across all centers combined. A full waiver of informed consent was granted, since the study extracts records from a de-identified repository with no patient contact.

