## Supplementary Material for "Automated, Component-Level Assessment of a National Inpatient Diabetes Guideline: Adherence and Its Association with Hospitalization Complications Across Six Hospitals"

This supplement gives the data-preparation detail, operational definitions, and model summary supporting the main text. Appendix S1 covers extraction, concept curation, derived concepts, the modified Charlson comorbidity index, threshold acquisition, score aggregation, and software. Appendix S2 defines the ten complications. Appendix S3 defines the fourteen assessed guideline recommendations. Table S2 summarizes the bias-reduced regression models. The final section gives an exploratory description of daily adherence-score trajectories.

### S1 Data Preparation, Knowledge Acquisition, and Software

**Source and unified tables.** The extraction consumed the OMOP tables `person`, `visit_occurrence`, `visit_detail`, `death`, `condition_occurrence`, `observation`, `procedure_occurrence`, `device_exposure`, `drug_exposure`, and `measurement`, and reorganized them into five unified tables: *visits*, *visit types*, *clinical events* (conditions, observations, procedures, and device exposures under a common schema), *drug exposures*, and *measurements*. The only enrichments applied were the attachment of Anatomical Therapeutic Chemical codes to drug exposures, where available, the harmonization of column types, and the closing of intervals whose end timestamp was missing or not later than their start one second after their start, so that point observations stored under an interval schema carry an explicit rather than a degenerate duration. Dimension tables enumerating the distinct clinical, measurement, and drug concepts, together with their units and administration routes, were derived from the unified tables and used to support concept curation.

**Concept curation.** Candidate codes for each target concept were surfaced by descriptive analytics over the dimension tables, ranked by frequency of occurrence and by apparent relevance to the concept, and then accepted or rejected by the clinical domain expert. The resulting indices define both the concepts included in the abstraction input and the departmental and diagnostic exclusions applied in the cohort definition. Exclusions were applied through these curated indices rather than inline rules, so that each exclusion is recorded once alongside the concept vocabulary and applied uniformly across the pipeline. The curated indices, rather than the exploratory analytics used to generate the candidate lists, are the reproducible artifact of this step, and are provided with this supplement.

**Derived concepts.** Body mass index was computed from paired height and weight measurements where not recorded directly. Ketoacidosis events were derived from combinations of pH, bicarbonate, and intravenous insulin administration, as described in Appendix S2.

**Modified Charlson comorbidity index.** The score is a weighted sum of ten chronic condition flags ascertained from the first 48 hours of admission: malignancy, chronic kidney disease, and chronic liver disease weighted 2; and cardiac disease, chronic respiratory disease, diabetes, dementia, alcohol misuse, anemia, and recurrent falls weighted 1. One additional point is added per decade of age above 50. Categories absent from the original Charlson instrument reflect the condition groupings that could be resolved from the unmapped source vocabulary during curation.

**Threshold acquisition.** Discretization thresholds, relevance durations, and trend limits were acquired in two passes. One-dimensional $k$-means and kernel density estimation over each concept's value distribution proposed candidate boundaries; these were then reconciled against published reference ranges and the definitions in the national guideline, which were authoritative, with the data-driven candidates used only to resolve cases where the literature gives a range rather than a value. Concepts with too many distinct value combinations for in-engine discretization, such as corticosteroid preparations at varying doses, were pre-discretized during data preparation.

**Aggregation of adherence scores.** The admission is the unit of analysis throughout. A patient may contribute several admissions, and patient-level grouping is applied wherever the statistics require it, as in the clustered standard errors of the mortality model. Compliance scores were aggregated in stages designed so that neither frequently-evaluated patterns nor frequently-evaluated days dominate the result. Scores were first averaged within each admission, pattern, compliance aspect, and calendar day; aspects were then collapsed within each admission, pattern, and day; days were then collapsed within each admission and pattern; and finally patterns were averaged with equal weight to give one overall adherence score per admission. Component-level analyses use the corresponding pattern-level and aspect-level aggregates.

**Software.** All processing and analysis were performed in Python 3.13. The extraction and cohort-construction pipeline used `dask` and `pyarrow` over Parquet storage, with `pandas` and `numpy` for in-memory operations; empirical threshold discovery used $k$-means clustering and kernel density estimation from `scikit-learn` with peak detection from `scipy`. Guideline adherence was computed using the Mediator abstraction engine[1], which validates the knowledge base against its XML schema before evaluating the abstractions. Statistical analysis used `statsmodels` for the logistic, propensity-weighted, and Welch comparisons, together with its Benjamini–Hochberg implementation for multiplicity correction. Bias-reduced logistic regression was implemented directly, following Firth's iteratively reweighted least-squares formulation with a Jeffreys-prior penalty and standard errors taken from the penalized information matrix; it is released with the analysis code rather than drawn from a library.

### S2 Operational Definitions of Complications

Definitions are given in the order used in the main-text tables. The glycemic endpoints and acute kidney injury are derived within the knowledge base from the corresponding measurements, while ketoacidosis, infection, hyperosmolality and myocardial infarction were computed during data preparation and supplied to the engine as primitive concepts.

**Thirty-day mortality.** Death recorded in the linked mortality data between 14 days before and 30 days after the recorded discharge datetime. The backward tolerance ensures that in-hospital deaths are captured where the death record precedes the administrative discharge timestamp.

**Hyperglycemia.** Either a single glucose measurement $\geq$250 mg/dL, or a sustained hyperglycemic state, defined as a glucose measurement $\geq$180 mg/dL whose immediately preceding measurement within the previous 24 hours was also $\geq$180 mg/dL. The sustained-state requirement prevents an isolated elevated reading from being counted as a complication.

**Severe hyperglycemia.** Any single glucose measurement $\geq$250 mg/dL. This endpoint overlaps by construction with hyperglycemia and is retained as a separate, more specific target.

**Hypoglycemia.** Either a single glucose measurement $\leq$54 mg/dL, or a sustained hypoglycemic state, defined as a glucose measurement $\leq$70 mg/dL whose immediately preceding measurement within the previous 24 hours was also $\leq$70 mg/dL.

**Severe hypoglycemia.** Any single glucose measurement $\leq$54 mg/dL.

**Inpatient acute kidney injury.** Laboratory-defined acute kidney injury of stage 2 or above, established either by a serum creatinine ratio $\geq$2.0 relative to the patient's baseline, or by an absolute serum creatinine $\geq$4.0 mg/dL. Stage 1 injury was deliberately excluded from the endpoint.

**Inpatient myocardial infarction.** Evidence of an ischemic cardiac event—ischemic heart disease, myocardial infarction, coronary intervention, cardiac catheterization, or revascularization—together with a troponin measurement of at least 600. The troponin requirement excludes coded cardiac history unaccompanied by biochemical evidence of acute injury.

**Hyperosmolality.** Emitted by either of two paths. The derived path implements a hyperosmolar-hyperglycemic-state rule: for each glucose measurement $\geq$600 mg/dL, the nearest serum sodium measurement within $\pm$6 hours is located and the effective osmolality computed as $2 \times \text{Na} + \text{glucose}/18$; an event is emitted where this exceeds 320 mOsm/kg, provided no acidosis event occurs within $\pm$6 hours of the triggering glucose draw. The acidosis exclusion separates hyperosmolar states from diabetic ketoacidosis. The observation path additionally emits an event wherever hyperosmolality is directly coded.

**Ketoacidosis.** Emitted by either of two paths. The observation path emits an event wherever ketoacidosis is directly coded. The derived path requires the conjunction of a pH measurement $\leq$7.3, a bicarbonate measurement $\leq$10 mEq/L within 24 hours of it, and intravenous insulin administration within 6 hours of it—that is, biochemical acidosis together with the treatment response it implies.

**Infection.** Requires a physiological trigger, corroborating diagnostic evidence, and treatment, all within a single episode. The trigger is any one of: fever onset, defined as at least two body-temperature measurements $\geq$37.8°C where all temperatures in the preceding 48 hours were $\leq$37.7°C; leukocytosis, defined as a white-cell count $\geq$11 where counts in the preceding 48 hours lay between 4 and 11; leukopenia, defined as a white-cell count $\leq$4 under the same prior-stability condition; or neutropenia, defined as a neutrophil count $\leq$1.5 where preceding counts within 48 hours exceeded 1.5. Diagnostic evidence requires at least one blood culture, urine culture, or chest radiograph within $\pm$24 hours of the trigger, the latest such record within the window serving as the episode anchor. Treatment requires at least one antibiotic administration after that anchor and within 72 hours of the trigger. Triggers occurring within 72 hours of an already-accepted event are collapsed into it, so an admission may contribute multiple distinct episodes. The prior-stability condition attached to every trigger restricts the endpoint to infections arising during the hospitalization rather than present on admission.

### S3 Assessed Guideline Recommendations

Fourteen quality-assessment patterns were evaluated, corresponding to the components reported in the main text (Table S1). Each pattern defines an applicability condition, so that it is scored only for admissions and time points at which the underlying recommendation applies, and one or more compliance aspects. Three aspect types are used: *time*, whether a required action occurred within a required interval; *cyclic*, whether a recurring action occurred at the required frequency; and *value*, whether an administered dose met the required magnitude. Insulin administration after hyperglycemia is a limiting case of the time aspect: it is defined by an overlap relation rather than by a delay, so no distance separates anchor from action for a membership function to grade, and the constraint is satisfied on existence alone—scoring 1 where the action is present and 0 where it is not.

Each aspect is scored by a trapezoidal membership function, so the reported values are continuous on $[0, 1]$: full credit is given throughout the interval in which the recommendation is considered met, with linear partial credit on either side of it and zero beyond the outer bounds. Several trapezoids are deliberately open on one side—full credit at or above a required count, for instance, with no penalty for exceeding it—so that partial credit applies only where departing from the recommendation in that direction is meaningful. A recommendation carrying more than one aspect is reported under each aspect separately, which is why routine basal dosage appears twice below—once for the frequency of administration and once for the dose administered. The knowledge base additionally contains a metformin continuation pattern; it is defined for detection only and carries no compliance function, so it yields no score and is not among the fourteen assessed components.

Table S1: Assessed guideline recommendations and their operational definitions in the knowledge base.

| Recommendation | Aspect | Operational definition |
|---|---|---|
| Admission glucose monitoring | Cyclic | Glucose measured in each 24-hour window over the first days of admission: three days where diabetes is documented, two otherwise. Full credit at three or more measurements per window, decaying linearly to zero at none. |
| Routine glucose monitoring | Cyclic | Glucose measured in each 24-hour window throughout the admission (evaluated to 14 days). While glucose is in a hyperglycemic or hypoglycemic state, full credit is given at three or more measurements per window. Under routine conditions the trapezoid peaks at at least one measurement per window. |
| Glucose monitoring during corticosteroid therapy | Cyclic | Glucose measured in each 24-hour window over the first two days of corticosteroid treatment, with full credit at at least three measurements per window. |
| BMI measurement on admission | Time | Body mass index or weight recorded within 48 hours of admission, decaying linearly to zero at 72 hours; a measurement recorded up to six months before admission is carried forward as relevant. |
| Creatinine measurement on admission | Time | Serum creatinine measured within 48 hours of admission, decaying linearly to zero at 72 hours. Unlike BMI, no pre-admission measurement is carried forward, so the measurement must fall within the admission itself. |
| Antidiabetic discontinuation on admission | Time | Prescribed antidiabetic agents carrying high hypoglycemia risk discontinued during the first 72 hours of admission, with credit decreasing linearly to zero if an agent was administered during that time. |

Continued on next page

Table S1: Assessed guideline recommendations and their operational definitions in the knowledge base (continued).

| Recommendation | Aspect | Operational definition |
|---|---|---|
| SGLT2 discontinuation on admission | Time | Prescribed SGLT2 inhibitors discontinued during admission. Any in-hospital administration scores 0, and the absence of one scores 1, assuming the patient was treated with the agent at home. |
| Basal insulin continuation on admission | Time | Basal insulin administered within 36 hours of admission for patients established on basal insulin at home, decaying linearly to zero at 48 hours. |
| Insulin administration after hyperglycemia | Time | Any dose of reactive bolus or intravenous insulin administered overlapping a meal (2-hour window before or after the meal) while glucose is elevated. |
| Antidiabetic discontinuation after hypoglycemia | Time | Prescribed antidiabetic agents withheld following a hypoglycemic event, with credit increasing over 72 hours and full credit sustained thereafter. |
| Basal dose reduction after hypoglycemia | Value | Basal insulin dose reduced relative to the preceding dose following a hypoglycemic event, scored on the ratio of the new dose to the preceding one: any reduction receives full credit, declining to zero at an unchanged dose. Unlike the bolus pattern below, hypoglycemic events *not* followed by a basal dose within 24 hours are scored as compliant rather than excluded, following the convention that a withheld action satisfies the recommendation. |
| Bolus dose reduction after hypoglycemia | Value | Bolus insulin dose reduced following a hypoglycemic event, on the same dose ratio and membership function as the basal pattern above, but evaluated only where bolus insulin was actually administered within 24 hours of the event; events with no subsequent bolus are excluded from scoring rather than counted as compliant (assuming basal is routine and bolus is a reaction to an event). |
| Routine basal dosage (frequency) | Cyclic | Basal insulin administered in each 24-hour window through the first 14 days of admission, evaluated in windows where hyperglycemia is present, with full credit between one and four administrations per window and declining credit beyond that range. |
| Routine basal dosage (dose) | Value | Judged against body weight for patients not established on basal insulin at home: full credit between 0.2 and 0.6 units per kilogram, rising from zero below 0.2 and declining to zero at 1.0 units per kilogram, using a default weight of 81 kg where no weight measurement is available. For patients already receiving basal insulin at home, the established dose is accepted as given and is not weight-normalized, assuming medical staff reacted to this dose and might have increased the hospital dose due to tolerance. |

The differing treatment of unanswered anchors in the two dose-reduction patterns is deliberate and reflects how the two insulins are used. Basal insulin is given routinely, so a hypoglycemic event with no basal dose in the following 24 hours represents the dose having been withheld, which satisfies the recommendation and scores 1. Bolus insulin is given reactively, so the absence of a dose carries no information about whether a reduction was warranted, and those events are excluded from scoring rather than counted as compliant. This is the principal reason the two components report such different means (0.87 and 0.28) despite sharing a membership function.

### S4 Summary of the Bias-Reduced Regression Models

Table S2: Summary of the bias-reduced (Firth) regression models. Positive-event counts are shown for each observation-window definition. Retained features denote predictors that remained significant after false discovery rate correction and pruning of highly correlated features. Robust features remained statistically significant with the same effect direction across all three observation-window definitions. AKI, acute kidney injury; MI, myocardial infarction.

| Outcome | 24 h | 48 h | 72 h | Retained | Robust |
|---|---|---|---|---|---|
| Inpatient MI | 175 | 125 | 90 | 13 | 0 |
| 30-day mortality | 3130 | 3130 | 2793 | 34 | 32 |
| Hyperglycemia | 8565 | 2153 | 1135 | 13 | 0 |
| Hyperosmolarity | 75 | 58 | 45 | 9 | 0 |
| Hypoglycemia | 2437 | 1742 | 1234 | 24 | 0 |
| Infection | 898 | 475 | 291 | 12 | 2 |
| Ketoacidosis | 46 | 33 | 25 | 9 | 0 |
| Inpatient AKI | 1362 | 901 | 655 | 21 | 5 |
| Severe hyperglycemia | 10019 | 4328 | 2376 | 28 | 8 |
| Severe hypoglycemia | 1686 | 1207 | 851 | 22 | 0 |

### S5 Daily Trajectories of Adherence Scores

We examined whether admission-level adherence scores reflected consistently similar daily performance or were driven by a small number of low-scoring days. Among admission–pattern pairs with at least three scored days (226,568 pairs), 36.1% had a minimum daily score at least 90% of their mean, whereas 32.9% had a minimum below 60% of their mean, indicating substantial heterogeneity in within-admission stability. Both trajectory types occurred at every hospital, although their frequency varied; for insulin after hyperglycemia, the share with a low-scoring day ranged from 30% to 60% across hospitals. Length of stay had only weak inverse correlations with mean daily adherence (Spearman $\rho = -0.110$) and within-admission variability ($\rho = -0.064$); the latter suggests little relationship between stay length and score variability despite its small $P$ value in this large sample. Daily trajectories were therefore mixed: roughly one-third were nearly flat, while a similar share of admission–pattern averages concealed one or more low-scoring days.
